# Cortical activation in healthy young adults performing a verbal fluency task during gait: A near-infrared functional spectroscopy (f-NIRS) study

**DOI:** 10.64898/2026.05.12.26353003

**Authors:** Felipe Augusto dos Santos Mendes, Paloma Rodrigues da Silva, Denilson Feijoeiro Garcia, Milena Satie Miamoto, Rauisa Gonçalves Macena, Larissa Bitarães Rodrigues Santos, Luiza de Mattos Aranha, Gabriel Venas Santos, João Ricardo Sato, Maria Elisa Pimentel Piemonte

## Abstract

**BACKGROUND:** Dual-task walking requires simultaneous management of cognitive and motor demands and is associated with changes in gait and cortical activation. However, the relationship between task-related cortical recruitment and dual-task-related gait adjustments in healthy young adults remains unclear. This study aimed to investigate the effects of dual-tasking on gait performance and cortical activation, and to examine the association between changes in cortical activity and dual-task costs.

**METHODS:** This cross-sectional study included 33 healthy young adults. Participants performed three conditions: single-task walking, cognitive single-task (verbal fluency), and dual-task walking. Each condition was repeated 10 times using a repeated short-block design with randomized trial presentation. Gait performance was assessed using an instrumented walkway, and cortical activation was measured using functional near-infrared spectroscopy. Dual-task costs were calculated for gait and cognitive outcomes.

Statistical analysis included repeated-measures analysis of variance (ANOVA) and Wilcoxon signed-rank tests, with false discovery rate correction for multiple comparisons. Associations between changes in cortical activation and dual-task costs were examined using correlation analyses.

**RESULTS:** Dual-task walking resulted in significant changes in gait, including reduced speed, step and stride length, and increased base of support, stance, and double support (all p < 0.05), while cognitive performance remained unchanged. Dual-tasking was associated with increased cortical activation in left prefrontal and motor-related regions. Greater increases in cortical activation were associated with lower dual-task costs across most gait parameters, with significant correlations observed in the left dorsolateral prefrontal cortex (r ≈ 0.42–0.47 for speed and stride length; p < 0.05). Double support showed a distinct pattern, suggesting a specific temporal adjustment within the gait cycle.

**CONCLUSIONS:** Dual-task walking in young adults is associated with coordinated behavioral and cortical adaptations. Increased cortical recruitment is linked to reduced motor interference, suggesting that broader engagement of cortical networks may contribute to performance under cognitive-motor load.

## INTRODUCTION

Gait has traditionally been described as an automatic motor behavior supported predominantly by spinal and subcortical mechanisms. Contemporary models, however, indicate that efficient locomotion emerges from the dynamic integration of spinal, subcortical, and cortical networks, particularly when task demands increase or environmental conditions require adaptation. In this framework, automaticity does not imply the absence of cortical involvement, but rather the ability to maintain locomotor performance with minimal reliance on executive control [1,2,3].

Dual-task paradigms have been widely used to probe the limits of gait automaticity and to investigate how cognitive and motor resources are allocated under competing demands. When a secondary cognitive task is introduced during walking, measurable changes in gait performance are often observed, including reductions in speed and step length and increases in parameters associated with stability, such as double support time and base of support [4]. These changes are typically interpreted as reflecting cognitive motor interference but may also represent adaptive strategies aimed at preserving safety under constrained attentional resources [5].

Importantly, dual-task walking is also associated with measurable changes in cortical activation. Studies using functional near-infrared spectroscopy (fNIRS) have consistently shown greater involvement of prefrontal and premotor regions while walking under cognitive load [6,7]. However, the functional significance of this response remains unresolved. While some studies interpret increased prefrontal recruitment as compensatory, supporting the maintenance of performance [8], others suggest that it may reflect increased processing cost or reduced neural efficiency, particularly when greater activation is associated with poorer motor outcomes [7,9].

This ambiguity is particularly evident in studies involving healthy young adults. While some investigations report minimal effects of dual-task on gait performance in this population, suggesting high motor-cognitive efficiency [4], others demonstrate measurable interference depending on task complexity and attentional demands [10]. Increased prefrontal activation during dual-task gait has been observed in healthy young adults [11,12]. Yet, whether this response reflects efficient adaptation, increased processing cost, or both remains unresolved. Evidence suggests that greater cortical recruitment may accompany maintained performance in some contexts [8], whereas in others more demanding dual-task conditions lead to both increased prefrontal activation and greater gait interference [10], reinforcing the task-dependent nature of these effects.

Taken together, these findings indicate that the effects of dual tasking on gait and cortical activation in young adults are not fully understood. In particular, it remains unclear how task-related changes in cortical activity relate to domain-specific changes in gait performance, and whether increased activation reflects beneficial adaptation or increased neural cost. Addressing this question is important not only for advancing our understanding of locomotor control in healthy individuals, but also for informing translational applications. The growing use of non-invasive brain stimulation targeting frontal and motor cortical regions to improve gait and cognitive–motor function highlights the need to better characterize the functional role of these regions under cognitive–motor load. Characterizing task-related patterns of cortical recruitment and their associations with behavioral outcomes in healthy young adults may provide a reference framework for interpreting similar findings in aging and neurological populations and for informing future therapeutic strategies.

Therefore, the aim of the present study was to investigate how dual-task walking influences gait performance and cortical activation patterns in healthy young adults, and to examine the associations between task-related changes in brain activity and dual-task costs across different gait domains.

## MATERIALS AND METHODS

### Study Design

This is an observational, cross-sectional study that followed all the guidelines of the Strengthening the Reporting of Observational Studies in Epidemiology (STROBE) Statement and recommendations of consensus guide to using functional near-infrared spectroscopy in posture and gait research [13].

All experimental procedures were standardized and can be replicated based on the detailed protocol described.

### Ethics statement

The study was approved by the Research Ethics Committee of the Hospital das Clínicas, Faculty of Medicine, University of São Paulo (CAAE: 67388816.2.0000.0065; Approval number 6.913.344).

### Setting

This study was conducted in the research laboratory of REDE AMPARO, part of CEPID NEUROMAT (www.neuromat.numec.prp.usp.br). The laboratory was accessible, flat, isolated from excessive noise, and had controlled temperature and lighting.

### Participants

A convenience sample was recruited through REDE AMPARO (http://www.amparo.numec.prp.usp.br). Eligible participants were young adults aged 20–30 years.

Inclusion criteria were: willingness to participate, absence of cognitive impairment (Montreal Cognitive Assessment [MoCA] score > 26) [14], absence of depressive symptoms (Beck Depression Inventory-II [BDI-II] score < 13) [15], and ability to walk independently.

Exclusion criteria included diagnosed neurological, musculoskeletal, or cardiorespiratory disorders; gait impairment indicated by a Dynamic Gait Index (DGI) score < 20 [16]; and uncorrected visual or auditory deficits that could compromise task performance.

### Sample Size

The sample size calculation was performed using G*Power 3.1.9.7. The selected parameters included F-tests, Anova: Repeated measures, within factors and a priori power analysis, with an alpha level of 0.05, power of 0.95, and an ηp2 set at 0.190 (related effect size of 0.48). The effect size was adopted based on the findings of Dong et al. (2025), which utilized a similar evaluation method, outcome measures, and study population to the present study, being determined based on the difference found in premotor and supplementary motor area cortical activation between single and dual-task gait conditions [17]. As a result, a total sample of 16 participants was calculated. Considering a 20% dropout rate, a minimal sample size of 20 participants was determined for this study.

### Experimental Design and task paradigm

All procedures were conducted in person. Participants were recruited via internet, phone, and social media platforms of Rede AMPARO and invited to attend an evaluation session at the Rede AMPARO–NEUROMAT research laboratory. After receiving detailed information about the study, those who agreed to participate signed an informed consent form.

This cross-sectional study involved a single individualized session conducted by a trained physiotherapist, lasting approximately one hour. Initially, participants completed a structured demographic and clinical assessment, including sex, age, years of education, cognitive performance (MoCA), and depressive symptoms (BDI) [14,15].

Cortical activity was then assessed using fNIRS during three experimental conditions: (i) single-task walking at a self-selected pace, (ii) cognitive single-task involving a verbal fluency task performed in a standing position, and (iii) dual-task walking combining gait and verbal fluency.

The conditions were presented in a randomized order across trials, with randomization applied within each block to minimize potential order and learning effects. Each condition was repeated ten times. Each trial lasted 10 seconds and was followed by a 30-second interval, including rest, repositioning, and preparation phases. The total duration of the experimental protocol was approximately 20 minutes. A schematic representation of the protocol is shown in

### Task-conditions

#### Single-Task Walking Condition

In the single-task walking condition, participants walked at a self-selected habitual speed along the walkway for 10 seconds, without any additional cognitive demand. This condition was designed to assess cortical activity associated with locomotion.

#### Cognitive Single Task Condition

In the cognitive single-task condition, participants performed a phonemic verbal fluency task while standing. At the beginning of each trial, a distinct target letter was presented, varying across trials to minimize repetition and learning effects. Participants were instructed to generate as many words as possible within a 10-second interval.

Verbal fluency task was selected as the secondary cognitive task due to its high cognitive and executive demand and its extensive use in dual-task paradigms. Verbal fluency task requires continuous lexical retrieval, semantic memory access, and executive monitoring, engaging distributed cortical networks, particularly within left frontal regions, including the dorsolateral prefrontal cortex and inferior frontal gyrus [18]. In addition, tasks with higher executive demand have been shown to induce greater cognitive-motor interference and increased prefrontal activation during walking compared to less demanding paradigms [10,19]. These characteristics make verbal fluency a suitable paradigm to probe the limits of gait automaticity and to examine cortical resource allocation under cognitively demanding conditions.

In standard neuropsychological assessments, verbal fluency tasks are typically administered over 60-second intervals. In the present study, each trial lasted 10 seconds and was repeated 10 times per condition. This adaptation was implemented to optimize compatibility with the block-design structure required for fNIRS data acquisition, allowing repeated measurement of cortical hemodynamic responses while minimizing fatigue and motion-related artifacts.

#### Dual-Task Walking Condition

In the dual-task walking condition, participants performed the phonemic verbal fluency task while walking at a self-selected speed. At the beginning of each trial, a new target letter was randomly selected from the same predefined set of letters used in the cognitive single-task condition. The order of letter presentation was independently randomized for both conditions to minimize potential learning effects and order-related biases. Participants were not instructed to prioritize either the motor or the cognitive task, allowing for natural allocation of attentional resources between gait and verbal performance.

#### Outcomes measures Gait performance

Gait performance was assessed using the GAITRite system (CIR Systems Inc., Franklin, NJ, USA), a pressure-sensitive walkway widely used for spatiotemporal gait analysis [20,21]. Participants walked at a self-selected habitual speed under single-task and dual-task conditions.

Spatiotemporal gait parameters, including gait speed, step length, stride length, step time, base of support, double support, and stance phase, were automatically recorded by the system’s embedded sensors and processed using the GAITRite software.

For each condition, gait parameters were averaged across trials to obtain representative values. fNIRS and gait data were acquired simultaneously, ensuring temporal correspondence between neurophysiological and motor measures.

#### Cortical activity

Cortical activity was indirectly assessed using functional near-infrared spectroscopy (fNIRS), a noninvasive neuroimaging technique that measures changes in oxygenated (HbO) and deoxygenated hemoglobin (HbR) concentrations, providing an indirect index of neuronal activity [22]. A wearable, portable, wireless NIRSport2 continuous-wave system (NIRx Medizintechnik GmbH, Berlin, Germany) was used, employing dual-wavelength illumination (760 and 850 nm) to quantify temporal variations in HbO and HbR.

A custom 66-channel probe configuration, consisting of 16 light sources and 30 detectors with an interoptode distance of approximately 30 mm, was mounted on an adjustable cap and positioned according to the international 10–20 EEG system. The montage was centered at Cz to ensure consistent positioning across participants and provided coverage of bilateral prefrontal and premotor regions (including Fp1, Fp2, F3, and F4), as illustrated in Figure 2. Sixteen additional short-separation channels (∼15 mm) were included to capture superficial hemodynamic signals for subsequent regression of extracerebral components [23].

**Figure 1.**
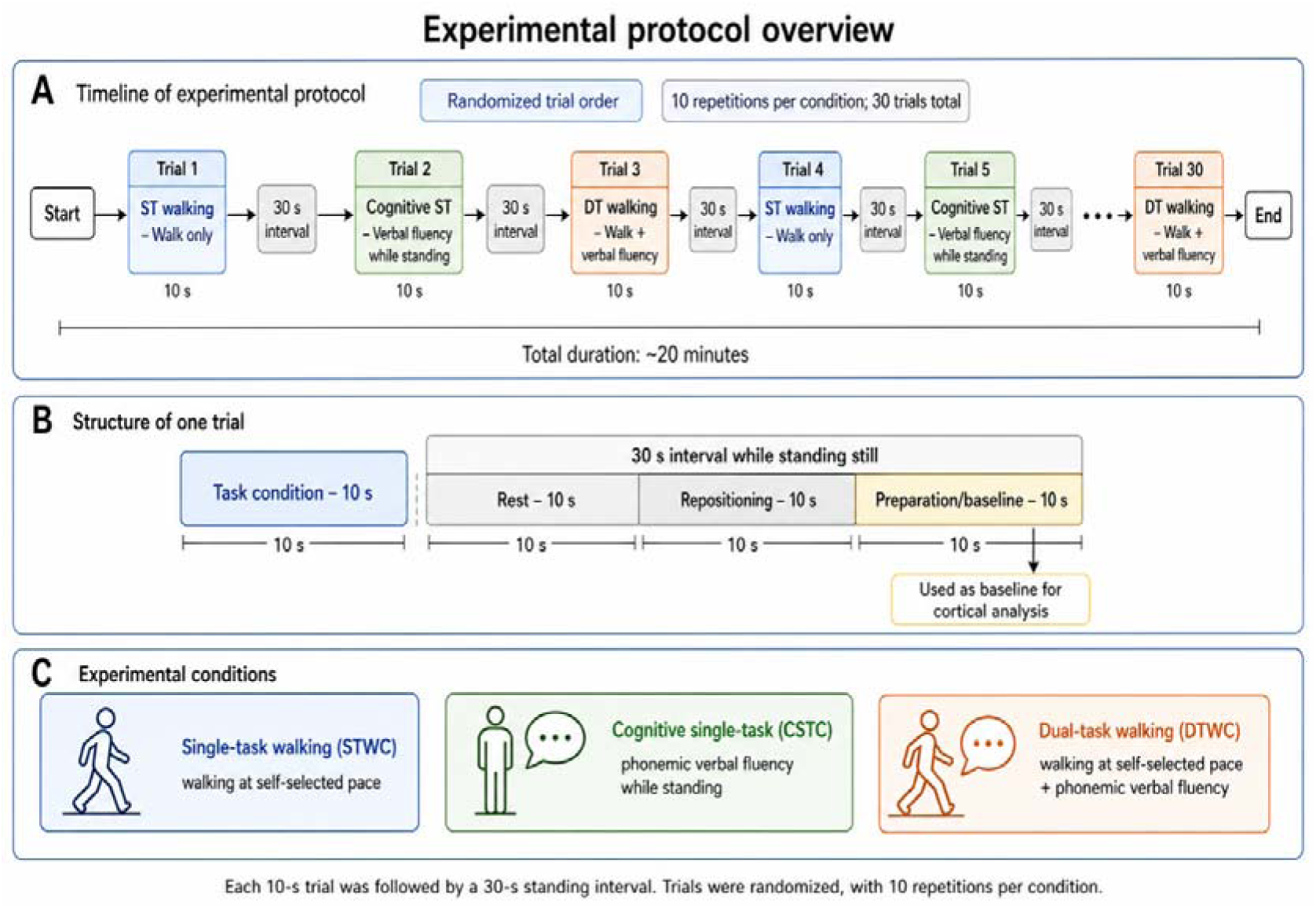
Experimental protocol overview. (A) Timeline of the experimental protocol. Participants performed 30 trials presented in randomized order, including 10 repetitions of each condition: single-task walking (STWC), cognitive single-task (CSTC), and dual-task walking (DTWC). Each trial lasted 10 seconds and was followed by a 30-second interval while standing still. The total duration of the protocol was approximately 20 minutes. (B) Structure of one trial. Each trial consisted of a 10-second task condition followed by a 30-second interval divided into three phases: rest (10 s), repositioning (10 s), and preparation (10 s). The final 10-second preparation phase was used as the baseline for cortical activity analysis. (C) Experimental conditions. STWC involved walking at a self-selected pace; CSTC consisted of a phonemic verbal fluency task performed while standing; and DTWC combined walking at a self-selected pace with the verbal fluency task.

**Figure 2.**
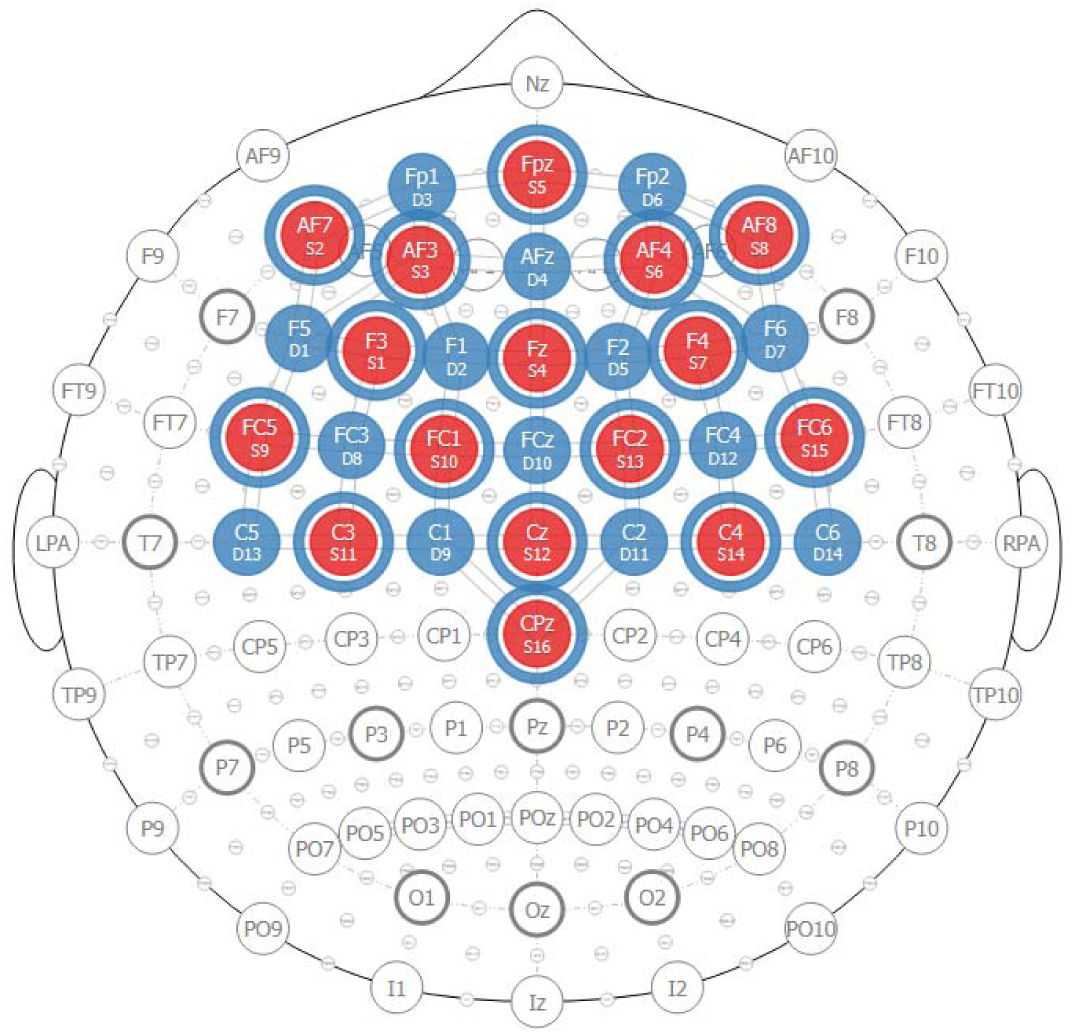
Schematic representation of the fNIRS analysis protocol. Red circles: Sources; Red circles circled in blue: Sources with short-distances detectors; Blue circles: regular detectors.

Data were acquired at a sampling frequency of 5.1 Hz and recorded using Aurora software (v.2021.4, NIRx Medical Technologies LLC, USA). The optode array was connected to a portable acquisition unit worn in a backpack, allowing data collection during walking. To minimize interference from ambient light, the cap was covered with a light-blocking layer during acquisition. System calibration was performed prior to data collection with participants in an upright standing position.

Raw optical signals were converted to changes in HbO and HbR concentrations using the modified Beer-Lambert law. Preprocessing included filtering and procedures to reduce physiological and motion-related artifacts (e.g., cardiac and respiratory components), as recommended in prior fNIRS studies [24].

Hemodynamic responses were analyzed using a general linear model. Task conditions were modeled as repeated 10-second blocks corresponding to the experimental paradigm, and condition-specific beta coefficients were estimated for each channel. Group-level analyses were conducted using individual beta values, accounting for within-subject structure and temporal autocorrelation.

For all analyses, HbO signals were used as the primary outcome measure due to their higher signal-to-noise ratio and greater sensitivity to task-related cortical activation compared to HbR [6,7].

#### Cognitive Performance Assessment

Cognitive performance was assessed using the number of correctly generated words during the phonemic verbal fluency task. Responses were recorded for each trial and subsequently scored offline according to the criteria described above, excluding proper nouns, repeated words, numerical responses, and morphological variants of previously generated words.

For each condition, the total number of valid words was averaged across trials to obtain a representative measure of cognitive performance. This outcome was used to quantify verbal fluency performance under cognitive single-task and dual-task conditions.

#### Cortical Activity Evaluation Across Task Conditions

Cortical hemodynamic responses were analyzed using a block-design approach aligned with the experimental protocol. Task-related activation was estimated relative to the baseline defined as the final 10-second period of each inter-block interval.

fNIRS and gait data were acquired simultaneously during all task conditions, ensuring temporal alignment between neurophysiological, motor, and cognitive measures.

## Statistical Analysis

Normality was assessed based on the distribution of paired differences between dual-task and single-task conditions using the Shapiro-Wilk test. For behavioral outcomes, paired differences between the two relevant conditions were examined. Gait outcomes were compared between single-task walking and dual-task walking, whereas cognitive performance was compared between cognitive single-task and dual-task conditions. Variables with approximately normal paired differences were analyzed using one-way repeated-measures ANOVA with two within-subject levels, while variables with non-normal paired differences were analyzed using Wilcoxon signed-rank tests.

To account for multiple comparisons across gait parameters, p-values were adjusted using the Benjamini-Hochberg false discovery rate procedure as the primary correction method. A Bonferroni correction was additionally applied as a conservative sensitivity analysis.

Dual-task costs were calculated for each outcome as (dual-task - single-task) / single-task.

Cortical hemodynamic responses were analyzed using one-way repeated-measures ANOVA, with condition (single-task walking, cognitive single-task, and dual-task walking) as a within-subject factor. When significant main effects were observed, Bonferroni-adjusted post hoc comparisons were performed.

Changes in cortical activation were calculated as ΔHbO (dual-task minus single-task). Associations between ΔHbO and dual-task gait and cognitive costs were examined using Pearson correlation coefficients when assumptions of normality were met and Spearman rank correlations otherwise.

Given the homogeneous characteristics of the sample (young age, high educational level, and preserved cognitive and motor function), no additional covariates were included in the statistical models.

No missing data was observed for gait or cognitive outcomes. For fNIRS data, all channels were retained for analysis as no predefined criteria for channel exclusion were applied, and signal quality was deemed acceptable across participants.

All analyses were performed using SPSS version 20 (IBM Corp., Armonk, NY, USA), with statistical significance set at α = 0.05.

### Potential sources of bias

Several strategies were implemented to minimize potential sources of bias. The randomized presentation of task conditions across trials reduced order and learning effects. The use of repeated trials and baseline periods aimed to improve signal stability and reduce variability in fNIRS measurements. In addition, standardized instructions were provided across all conditions, and no task prioritization was imposed, allowing natural allocation of attentional resources. The homogeneous sample reduced variability related to demographic and clinical factors.

## RESULTS

### Sample characteristics

A total of 36 individuals were assessed for eligibility. Three participants declined participation due to scheduling incompatibility. No participants were excluded based on eligibility criteria. A final sample of 33 participants was included, all of whom completed the protocol and were included in the analysis, as shown in Figure 3.

**Figure 3:**
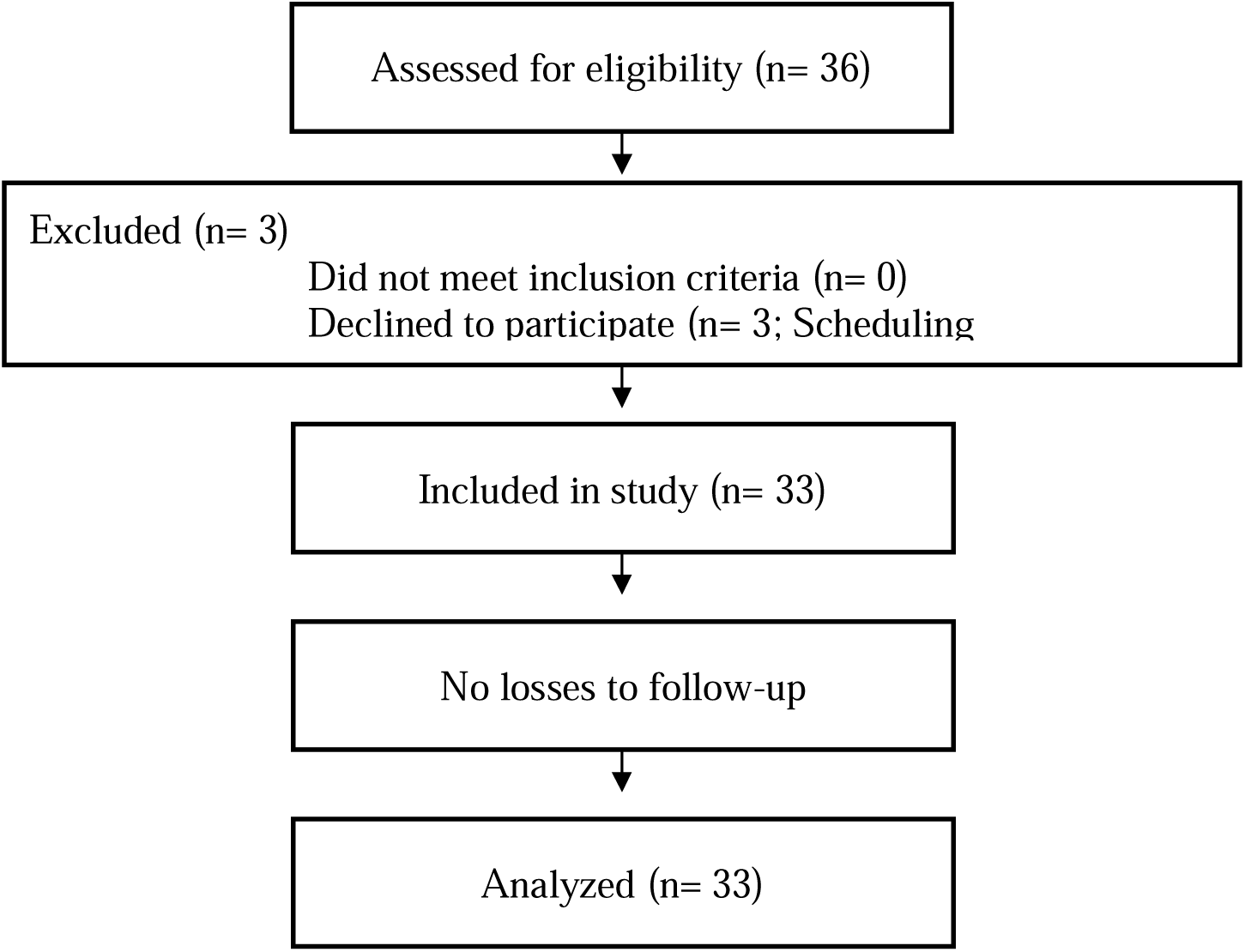
Participant flow diagram.

The sample characteristics are presented in Table 1. Participants were young adults with a high educational level and preserved cognitive, mood, and motor performance.

**Table 1.**
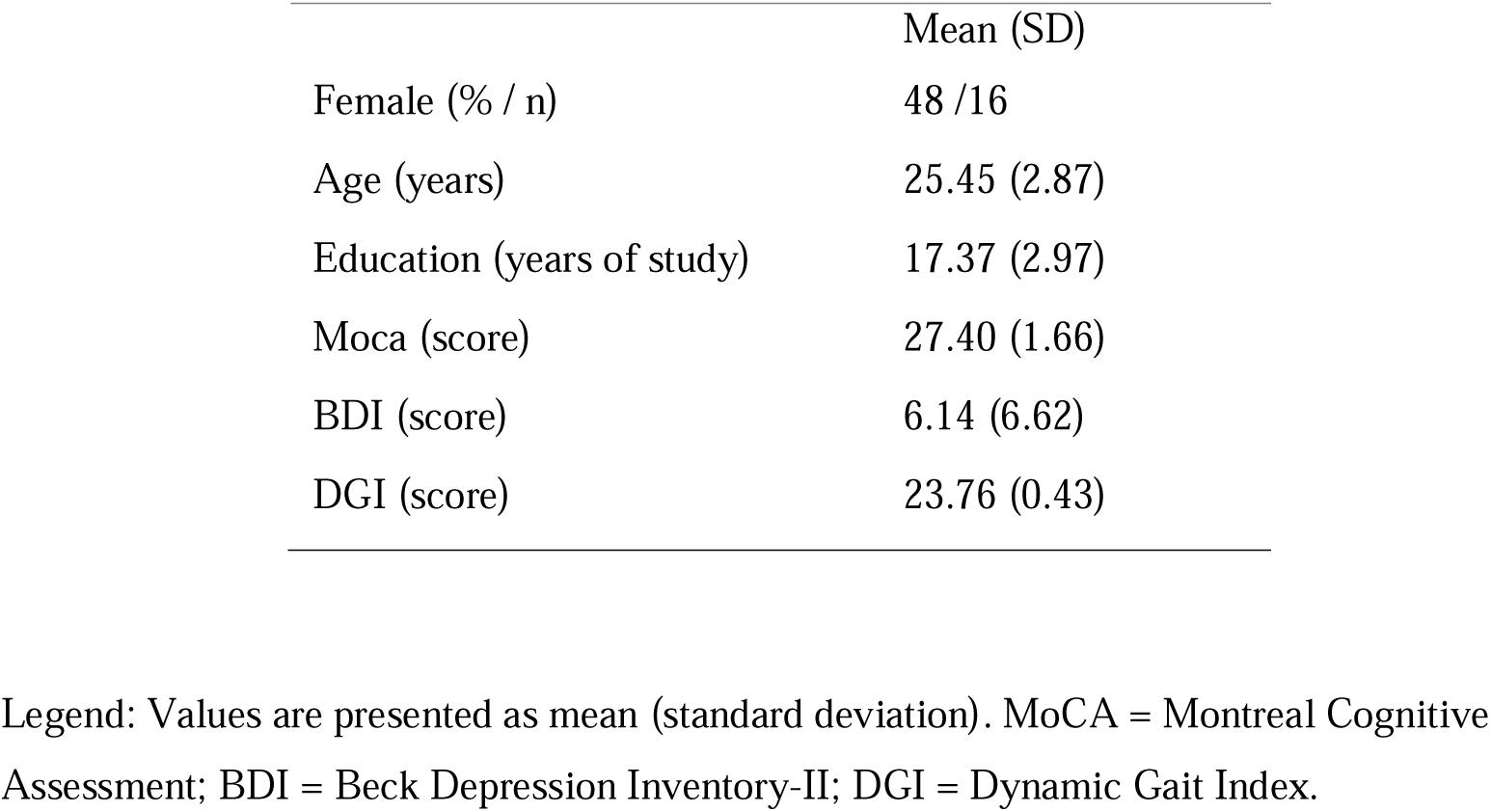
Demographic data of participants (n = 33)

### Gait and cognitive performance

After false discovery rate correction, significant differences between conditions were observed for gait velocity, step time, step length, stride length, H–H base of support, single support, double support, and stance. Compared to single-task walking, dual-task walking was characterized by lower velocity, longer step time, shorter and stride lengths, wider base of support, reduced single support, increased double support, and increased stance.

In a conservative sensitivity analysis using Bonferroni correction, all of these effects remained significant except for step time, which was significant after false discovery rate correction but not after Bonferroni adjustment. Swing phase was significant only in the unadjusted analysis and did not remain significant after either correction method.

No significant differences were observed for cycle time, toe in/out, or cognitive performance (mean number of words) between single-task and dual-task conditions.

### Cortical activation across conditions

Cortical activation levels (oxygenated hemoglobin, HbO) across conditions are presented in Figure 4. One-way repeated-measures ANOVA revealed significant effects of condition in the left dorsolateral prefrontal cortex, premotor cortex, and supplementary motor area. No significant effects were observed in the other cortical regions evaluated. Bonferroni-adjusted post hoc comparisons showed greater activation in these regions during dual-task walking compared to single-task walking (p < 0.006). No significant differences were observed between dual-task and the cognitive single-task condition.

**Figure 4.**
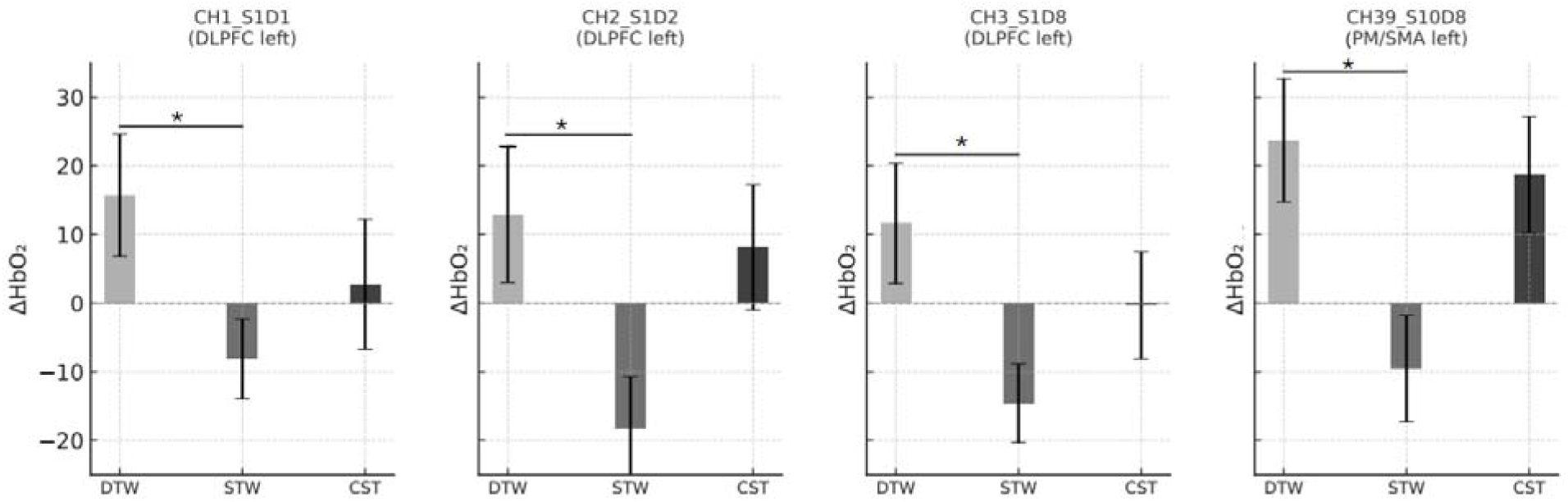
Comparisons of oxygenated Hemoglobin (μmol/L) variations in dual and single tasks. ΔHbO[ – Oxyhemoglobin variations (means and standard errors); *p<0.006 (Bonferroni post hoc correction), **p<0.001; DLPFC - Dorsolateral prefrontal cortex; PM_SMA - Premotor and supplementary motor cortex; DTW - Dual-task walking; STW - Single-task walking; CST - Single cognitive task.

### Associations between cortical activation and dual-task costs

Correlation coefficients between changes in cortical activation (ΔHbO) and dual-task gait costs are presented in Table 2.

**Table 2:** Relationship between cortical hemodynamics responses variation under ST and DT conditions and DT gait costs.

| DT gait costs | Cortical areas |  |  |  |
| --- | --- | --- | --- | --- |
|  | CH1_S1D | CH2_S2D | CH3_S1D | CH39_S10D |
|  | 1 DLPFC | 2 DLPFC | 8 DLPFC | 8 PM_SMA |

|  | (left) | (left) | (left) | (Left) |
| --- | --- | --- | --- | --- |
| Speed | – | – | 0.42*<br>(0.02) | – |
| Steplength | – | – | – | – |
| Stride length | – | – | 0.47<br>(0.01) | – |
| Base of support | -0.55<br>($<0.01$) | -0.40<br>(0.02) | -0.43<br>(0.01) | -0.47<br>(0.01) |
| Double Support | 0.38*<br>(0.04) | – | – | – |
| Stance | – | – | -0.60<br>($<0.01$) | -0.39<br>(0.03) |

Legend: DLPFC - Dorsolateral prefrontal cortex; PM\_SMA - Pre Motor and Supplementary Motor Cortex; DT - Dual task; \*Pearson correlations - nonsignificant.

Significant correlations were observed primarily in the left dorsolateral prefrontal cortex. In this region, ΔHbO showed positive correlations with the dual-task costs of gait speed and stride length, and negative correlations with the costs of base of support, double support, and stance. Considering that, for speed and stride length, more negative cost values reflect greater performance reduction, and for base of support, double support, and stance, more positive values reflect increased reliance on support-related strategies, these correlation patterns consistently indicate that greater increases in cortical activation were associated with lower dual-task interference across gait parameters.

Premotor cortical regions also demonstrated significant negative correlations with the dual-task costs of base of support, double support, and stance. Considering that more positive cost values for these parameters reflect increased reliance on support-related strategies, these findings indicate that greater increases in premotor cortical activation were associated with reduced dual-task interference in gait stability-related measures.

No significant correlations were observed between cortical activation and the dual-task costs of step length or cognitive performance (verbal fluency task).

In summary, dual-tasking significantly impacted gait performance in young adults, leading to reduced speed and spatial parameters and increased temporal measures associated with gait support, while cognitive performance remained unchanged. At the neural level, dual-task walking was associated with increased activation in left prefrontal and motor-related cortical regions, including the dorsolateral prefrontal cortex, premotor cortex, and supplementary motor area. Furthermore, greater task-related increases in cortical activation were significantly associated with dual-task gait costs across multiple parameters.

## DISCUSSION

Although gait has traditionally been considered an automatic motor behavior, accumulating evidence indicates that it depends on dynamic cortical integration, particularly under cognitively demanding conditions. While dual-task interference on gait has been extensively described in older adults and neurological populations, less is known about how cognitively preserved young adults respond to dual-task demands, especially regarding the relationship between cortical activation and gait performance. The present study addressed this gap by examining both behavioral and neural responses to dual-task walking in a high-functioning young adult sample.

The findings showed that dual-tasking induced measurable changes in gait performance, even in this population, characterized by reduced speed and spatial parameters and increased support phases, while cognitive performance remained preserved. Importantly, these changes were accompanied by increased activation in frontal and motor-related cortical regions, and greater cortical recruitment was predominantly associated with lower dual-task gait costs.

### Cortical recruitment and modulation of dual-task costs

The primary evidence emerging from this study is that increased cortical activation, particularly in the left dorsolateral prefrontal cortex, was associated with reduced dual-task costs across most gait parameters. This pattern suggests that cortical recruitment represents an adaptive mechanism supporting the concurrent management of motor and cognitive demands [7,8,11], rather than reflecting inefficiency or overload [6,13]. Individuals with greater activation exhibited better preservation of gait speed, stride length, base of support, and stance, indicating that increased neural engagement contributes to maintaining the overall organization of gait under dual-task conditions [8,12,17].

Notably, double support was the only parameter showing an association in the opposite direction. This finding requires careful interpretation. Double support represents the portion of the gait cycle during which both feet are simultaneously in contact with the ground and is a distinct temporal component of stance, complementary to single support; GAITRite-derived gait analyses commonly report these temporal parameters separately, including stance, single support, double support, and swing phases [20]. Therefore, changes in double support should be interpreted as a redistribution of temporal phases within the gait cycle, particularly during weight transfer between limbs, rather than as a standalone marker of global gait deterioration. Increased double support is commonly observed as part of a more cautious gait pattern, often alongside slower gait speed, shorter steps, and wider step width/base of support [25]. However, double support also has a specific biomechanical role in step-to-step transition and mechanical stabilization during walking, supporting the interpretation that this parameter may capture a localized temporal adjustment rather than a generalized decline in gait organization [26]. Together, these findings indicate that increased cortical recruitment was predominantly associated with reduced dual-task costs in the overall organization of gait, while the behavior of double support may reflect a more specific temporal adjustment within the locomotor pattern.

Beyond the prefrontal cortex, significant associations were also observed in premotor and supplementary motor regions, particularly in relation to stability-related gait parameters. In these areas, increased cortical activation was consistently associated with reduced dual-task costs in base of support, double support, and stance, reinforcing the pattern observed in the dorsolateral prefrontal cortex. These findings are consistent with the role of premotor and supplementary motor areas in movement planning, temporal organization, and postural preparation [27,28,29,30].

This pattern provides important insight into the neural mechanisms underlying dual-task performance. If gait control were simply shifted toward a more automatic mode to free cortical resources for the cognitive task, a reduction in motor cortical involvement would be expected [1,2,3]. However, the observed increase in premotor and supplementary motor area activation suggests the opposite: dual-tasking appears to reduce gait automaticity, requiring additional recruitment of regions involved in motor planning and coordination. This interpretation is consistent with evidence that highly automatic movements are associated with more efficient and localized neural processing, whereas reduced automaticity increases dependence on distributed fronto-motor networks [5,6,11].

An additional factor that must be considered is that the verbal fluency task is not purely cognitive. In addition to lexical retrieval and executive processing, it involves overt speech production, which depends on motor planning and articulatory control [31]. Thus, the dual-task condition required the simultaneous execution of two motor behaviors with partially automatic characteristics, locomotion and speech production. The increased activation observed in premotor and supplementary motor areas may therefore reflect not only cognitive–motor interference but also the need to coordinate concurrent motor outputs, including gait, postural control, and speech articulation.

In addition, the predominantly left-lateralized pattern of activation observed in both prefrontal and motor-related regions may reflect the linguistic demands of the task.

Language processing and speech production are typically associated with left-hemisphere networks, including frontal and premotor areas, and increased recruitment in these regions during dual-task walking may therefore be influenced by the verbal nature of the secondary task [10,18,31]. Although this lateralization should be interpreted with caution, it is consistent with the hypothesis that task-specific demands contribute to the observed cortical activation profile.

Taken together, these findings suggest that dual-task walking engages a distributed cortical network involving both executive and motor planning regions. Within this network, prefrontal areas may support resource allocation and task management, whereas premotor and supplementary motor regions contribute to the coordination and temporal organization of movement, particularly under conditions requiring the simultaneous control of gait and speech.

### Dual-task effects: gait reorganization with preserved cognitive performance

A second key finding is that dual-tasking affected multiple gait parameters while cognitive performance remained stable. Participants exhibited reductions in gait speed and step and stride length, along with increases in base of support, stance, and double support, indicating a consistent reorganization of gait under cognitive load. Despite these changes, verbal fluency performance was preserved, suggesting that the dual-task condition did not compromise cognitive output.

This pattern is consistent with models of limited attentional capacity and dual-task interference, which propose that when tasks compete for shared neural resources, performance in one domain may be maintained at the expense of another [32]. In the present study, the preservation of cognitive performance alongside systematic changes in gait suggests a redistribution of resources favoring cognitive demands, resulting in adjustments in motor performance rather than cognitive decline. This interpretation is further supported by studies demonstrating that individuals may adopt different prioritization strategies under dual-task conditions, allocating attention preferentially to either cognitive or motor tasks depending on task demands [33].

Importantly, this should not be interpreted as a failure of motor control. Instead, the observed changes likely reflect an adaptive reorganization of gait to maintain functional stability under cognitive load, even in young adults with high cognitive reserves. This perspective aligns with frameworks of cognitive-motor interaction, which emphasize the dynamic interplay between executive control and motor performance during complex tasks [5,34,35].

From a broader perspective, cognitive resources are limited and must be dynamically allocated according to task demands [6,32]. Under such conditions, preserving cognitive processing, particularly functions related to environmental monitoring, decision-making, and response selection, may be prioritized, while motor output is flexibly adjusted to maintain overall task performance. This adaptive allocation becomes especially relevant in more complex or ecologically valid contexts, where cognitive demands may increase and influence locomotor behavior [10,36,37].

Within this framework, the changes observed in gait during dual-tasking should not be interpreted solely as deficits, but rather as part of a strategic allocation of resources that allows the maintenance of cognitive performance while preserving functional mobility.

## CLINICAL IMPLICATIONS

The present findings have direct implications for neurorehabilitation, particularly in the context of growing interest in interventions targeting cognitive-motor interactions during locomotion. The observation that dual-task walking engages a distributed cortical network, including prefrontal, premotor, and supplementary motor regions, suggests that these areas may play complementary roles in supporting gait under cognitively demanding conditions. From a clinical perspective, this reinforces the need to move beyond single-domain approaches and to incorporate dual-task paradigms into both assessment and intervention strategies.

The identification of region-specific patterns of activation associated with reduced dual-task costs also has potential relevance for neuromodulation approaches. With the increasing use of non-invasive brain stimulation techniques, such as transcranial magnetic stimulation and transcranial direct current stimulation, understanding the functional contribution of specific cortical regions during dual-task walking becomes critical. The present results suggest that the dorsolateral prefrontal cortex may be a key target for modulating executive aspects of dual-task performance, whereas premotor and supplementary motor areas may represent additional targets for improving the coordination and temporal organization of movement under cognitive load.

These considerations are particularly relevant for older adults and clinical populations, such as individuals with Parkinson’s disease or other neurological conditions, who often exhibit pronounced impairments in dual-task performance. In such populations, deficits may arise not only from reduced motor automaticity but also from altered cortical recruitment patterns.

Finally, the present findings support the use of ecologically valid assessment paradigms that integrate motor and cognitive demands, as these better reflect real-world functional challenges. Identifying individual patterns of cortical activation and dual-task cost profiles may help guide personalized rehabilitation strategies, including the selection of stimulation targets and the design of task-specific training protocols.

## STRENGTHS AND LIMITATIONS

This study presents several methodological strengths that contribute to the robustness and interpretability of the findings. First, the experimental design incorporated a high number of repetitions (10 trials per condition) within a block paradigm, combined with systematic baseline periods preceding each trial. This approach is particularly relevant for fNIRS, as it enhances signal reliability and increases the sensitivity to detect task-related hemodynamic contrasts that might not be captured in studies using fewer repetitions or less structured protocols.

Second, the choice of the verbal fluency task as the cognitive component proved to be appropriate for the study objectives. Despite the relatively short duration of each trial (10 seconds), the task was sufficiently demanding to induce measurable dual-task interference, as evidenced by consistent changes in gait parameters. This supports the use of brief, repeated cognitive challenges within fNIRS-compatible paradigms.

Third, the study combined a comprehensive assessment of cortical activation across multiple frontal and motor-related regions with a detailed analysis of gait performance. The use of an instrumented walkway allowed the extraction of multiple spatiotemporal parameters, providing a nuanced characterization of gait adjustments under dual-task conditions. In parallel, cognitive performance was continuously monitored, and baseline conditions were established for both motor and cognitive tasks. This design enabled the calculation of both motor and cognitive dual-task costs, offering an integrated perspective on cognitive–motor interaction.

Despite these strengths, some limitations should be considered. The relatively small sample size may limit the generalizability of the findings and the statistical power to detect more subtle effects, particularly in correlation analyses. In addition, the study included a homogeneous sample of young, highly educated individuals, which, while advantageous for internal validity, may restrict the applicability of the results to populations with different demographic or clinical characteristics.

Furthermore, the duration of each trial was shorter than that typically used in standard neuropsychological assessments of verbal fluency. Although this was necessary to ensure compatibility with the fNIRS block design, it may have influenced the sensitivity of the cognitive outcome measures.

## CONCLUSION

Dual-task walking in healthy young adults led to consistent adjustments in gait parameters accompanied by changes in cortical activation patterns, particularly in prefrontal and motor-related regions. Importantly, greater increases in cortical activation were associated with smaller dual-task–related changes in gait, indicating that broader cortical recruitment was linked to reduced motor impact of dual-tasking.

## DECLARATIONS

### Ethics approval and consent to participate

The study was approved by the Research Ethics Committee of the Hospital das Clínicas, Faculty of Medicine, University of São Paulo (CAAE: 67388816.2.0000.0065; Approval number 6.913.344). All participants were informed about the study procedures and objectives prior to participation and provided written informed consent before the assessments were conducted.

### Consent to publish

Prior to the beginning of the assessments all participants were informed about the research and signed a free informed consent form, authorizing the analysis and publication of the data obtained from their participation.

### Data availability

The datasets generated and/or analyzed during the current study are not publicly available but are available from the corresponding author on reasonable request.

### Competing interests

The authors declare that they have no competing interests.

## Funding

This study was funded by the São Paulo Research Foundation (FAPESP) through the Research, Innovation and Dissemination Center for Neuromathematics (CEPID NeuroMat, grant number 2013/07699-0).

Co-author PRS received individual support from FAPESP (grant number 2025/14403-7).

Co-author DFG received individual support from FAPESP (grant number 2024/16868-4).

Co-author MSM received individual support from FAPESP (grant number 2024/15842-1).

Co-author RGM received individual support from FAPESP (grant number 2024/15841-5).

Co-author GVS received individual support from FAPESP (grant number 2025/02885-7).

Co-author JRS received individual support from FAPESP (grant number 2023/18337-3; 2023/16997-6; 2023/02538-0).

## Authors’ contributions

MEPP and JRS designed the study. MEPP, JRS, PRS, LBRS and LMA designed the assessment protocol. FASM, PRS, LBRS, LMA, DFG, MSM, RGM, GVS and JRS collected the data and were involved in data processing and data analysis. Statistical analysis was performed by FASM, MEPP and JRS. The draft of the manuscript was prepared by FASM, PRS, DFG, and LMA, and was critically reviewed by FASM, MEPP and JRS.

## Acknowledgments

This paper is dedicated to the memory of Professor Antonio Galves, whose unwavering commitment to the CEPID project laid the foundation for a multidisciplinary research group destined to thrive for years to come.

The authors sincerely thank all participants for their collaboration and commitment, as well as the technical and administrative teams who provided essential support in carrying out the research activities. We also acknowledge the institutions involved for their continuous support in the development of this work.

## Notes

### Competing Interest Statement

The authors have declared no competing interest.

### Funding Statement

This study was funded by the Sao Paulo Research Foundation (FAPESP) through the Research, Innovation and Dissemination Center for Neuromathematics (CEPID NeuroMat, grant number 2013/07699-0).
Co-author PRS received individual support from FAPESP (grant number 2025/14403-7).
Co-author DFG received individual support from FAPESP (grant number 2024/16868-4).
Co-author MSM received individual support from FAPESP (grant number 2024/15842-1).
Co-author RGM received individual support from FAPESP (grant number 2024/15841-5).
Co-author GVS received individual support from FAPESP (grant number 2025/02885-7).
Co-author JRS received individual support from FAPESP (grant number 2023/18337-3; 2023/16997-6; 2023/02538-0).

### Author Declarations

Research Ethics Committee of the Hospital das Clinicas, Faculty of Medicine, University of Sao Paulo (CAAE: 67388816.2.0000.0065; Approval number 6.913.344).

